# Hybrid lexical-semantic retrieval over SNOMED CT: combining two retrieval paradigms to facilitate clinical data entry

**DOI:** 10.64898/2026.09.10.26362796

**Authors:** Alejandro Lopez Osornio, Anne Randorff Højen, Kai Kewley

## Abstract

**Objectives:** Searching SNOMED CT pits two retrieval technologies of opposite nature against each other. Deterministic lexical matching is precise and handles partial, out-of-order, as-typed input, but cannot cross a wording or language gap; learned semantic matching bridges paraphrase and language, but represents abbreviations and half-typed fragments poorly. Combining them naively lets the channel that is wrong for a given query degrade the one that is right, and the usual alternative, curating an additional local interface vocabulary, is labour-intensive to maintain. We ask whether the two paradigms can instead *coexist* in one retrieval flow over SNOMED CT’s own curated descriptions, and study the query interpretation, rank fusion and re-ranking that make this coexistence safe.

**Materials and Methods:** We built one open-source, reproducible implementation over the SNOMED CT International edition, combining order-independent multi-prefix lexical search, BioLORD-2023-M semantic embeddings, an optional cross-encoder re-ranker, and a local LLM for query normalization and entity extraction, with the two rankings fused by Reciprocal Rank Fusion and an optional hierarchy (descendant) filter. We measured search-only linking accuracy on 985 disease mentions from DisTEMIST (Spanish, zero-shot), ablating each optional component and each retrieval channel, and on 12,897 mentions from the SNOMED CT Entity Linking Challenge (English real-EHR discharge notes) under a field-scoped typeahead.

**Results:** On DisTEMIST the best and cheapest configuration, semantic search with re-ranking and no LLM pre-processing, placed the exact concept first for 57% of mentions (accuracy@1 = 0.57) and within the top-10 for 80% (recall@10 = 0.80). Channel ablation showed the two paradigms coexist without a trade-off: rank fusion followed by re-ranking keeps the on-task channel’s result and suppresses the off-task one. On the English real-EHR corpus a field-scoped typeahead reached the top-10 for 73% of mentions across all clinical domains (accuracy@1 0.52). Adding a contextual LLM re-ranker — a second implementation of the re-ranking step that reorders the retrieved candidates using the query’s context — raised accuracy@1 further, to 0.61 on DisTEMIST and 0.56 on the challenge. LLM query normalization, by contrast, slightly hurt clean, terminology-like input but rescued the messy shorthand of real EHR text, most of all when given the surrounding note context, so it is best applied selectively rather than to every query. Results are specific to the evaluated configuration, not an architecture-independent estimate.

**Conclusion:** Lexical precision and semantic flexibility can coexist over SNOMED CT’s maintained descriptions without trading one off against the other, secured by rank fusion and re-ranking rather than by choosing between the paradigms. This shifts maintenance from local phrase authoring toward governing and validating retrieval services rather than removing it, and may reduce additional local lexical curation where direct search is appropriate, while curated interface vocabularies retain a role in guided workflows.

## Background and significance

### The vocabulary challenge

Coding with SNOMED CT can be implemented effectively using its distributed content alone: its concepts carry curated, clinician-authored descriptions that a terminology server can retrieve directly. Some implementations, however, ask more of that search than the shipped descriptions and a user’s own query skill readily provide, and there an underlying mismatch surfaces. Reference terminologies are engineered for concept permanence, formal definition and computability (Cimino 1998), whereas clinicians use compact, variable and often idiosyncratic expressions. This vocabulary challenge is inherent to human communication: when people name the same thing, they choose the same word less than 20% of the time (Furnas et al. 1987).

The challenge becomes concrete at the point of *clinical data entry*, where a free-text expression must be encoded to a specific SNOMED CT concept. Traditionally this encoding is performed interactively by a clinician who types a term into a search box and selects a concept from the results, an act of iterative query refinement that depends on the user’s familiarity both with how the terminology is worded and with how the matching algorithm behaves (Rosenbloom et al. 2006). Increasingly, the same encoding is also performed automatically, by artificial-intelligence agents that extract clinical entities from narrative text and normalize each to a concept with no human in the loop. The two modes stress the terminology in different ways: an interactive user’s input varies unpredictably (from a complete term to a short, partial, as-typed fragment or abbreviation, refined keystroke by keystroke), whereas an automated agent tends to submit a complete, already-normalized phrase, often translated into a common working language. Both, however, must cross the same gap between spontaneous clinical expression and curated terminology, and a retrieval interface intended to facilitate data entry must therefore serve both the fragmentary, interactive query and the normalized, batch one over the same content.

Terminology developers and implementers have addressed this challenge through different strategies. Efficient lexical algorithms improve access to the descriptions already curated within the terminology. When that is insufficient, interface terminologies add locally curated expressions intended to anticipate how users may search. More recently, language models have made it possible to compute semantic similarity at query time instead. These approaches place the work of resolving the vocabulary mismatch, together with its corresponding maintenance burden, in different places: within the reference terminology, with the local implementer or in the retrieval system. Our hypothesis is twofold: that dynamic semantic matching can replace much of the additional local lexical curation while continuing to rely on SNOMED CT’s maintained descriptions; and that it can do so *alongside*, not instead of, algorithmic lexical search: the two coexisting so that each covers the queries the other cannot, without either degrading the other.

### Curated descriptions and algorithmic lexical search

SNOMED CT already includes substantial curated lexical content: its concepts are accompanied by clinician-authored descriptions, including preferred terms and synonyms (approximately 1.6M active descriptions for approximately 368k concepts in the 2024 International edition) (SNOMED International 2024). This content reflects two important development traditions. Clinical Terms Version 3 (CTV3, formerly the Read Codes) was explicitly user-led and stressed the inclusion of the natural terms used by clinicians (O’Neil et al. 1995). In parallel, SNOMED RT was developed through a collaborative process involving the College of American Pathologists and Kaiser Permanente’s Convergent Medical Terminology (CMT) team (Levy et al. 1998). The merger of CTV3 and SNOMED RT brought these traditions together in SNOMED CT. This is also the genealogy of the clinician-facing vocabulary embedded in SNOMED CT: a centrally curated interface resource distributed and maintained with the reference terminology, rather than separately by each implementation.

In routine use, modern terminology servers combine this lexical content with fast, responsive search. Part-word retrieval has a long lineage in UK clinical terminology systems: clinicians searched Read Codes and CTV3 through the NHS Clinical Terminology Browser by entering one or more words or part-words (Brown et al. 2003), and later tooling for Read-coded data formalized searches using multiple word-stubs occurring anywhere in a description (Olier et al. 2016). The *multi-prefix, any-order* technique used here follows this interaction pattern: users enter only the first characters of one or more words, and each fragment is matched against the beginning of a word in the description, regardless of word order. Thus, the fragments *myo inf* can retrieve *myocardial infarction*, and reversing them does not change the match. This tolerates incomplete words and natural variation in word order while preserving precision by requiring every entered prefix. Because inflected and derived forms often share their initial characters, it also provides lightweight morphological normalization across variations such as number and, in languages that mark it, gender. This benefit is language-dependent and does not amount to full lemmatization. Clinicians can start with only a few characters and refine them in response to the results until they reach the concept they intend. This simple interaction makes the user an active participant in resolving the vocabulary mismatch, without requiring every possible expression to be anticipated in advance.

### Additional local lexical curation

Some implementations, however, need to reduce that dependence on individual search skills. Adoption constraints, limited training opportunities or the demands of a particular workflow may call for more natural queries and more guided concept selection. The conventional response is an additional *interface terminology*: a “systematic collection of health care-related phrases that support clinicians’ entry of patient-related information” (Rosenbloom et al. 2006). Added to the descriptions distributed with SNOMED CT, these local vocabularies map further colloquial expressions, abbreviations and preferred terms to concepts in the reference terminology. In effect, an interface terminology strengthens the lexical channel: it supplies more surface forms for each concept so that more queries succeed by wording alone. Until the recent maturation of efficient semantic retrieval, this was essentially the only way to broaden coverage without depending on the user’s search skill: adding a second, semantic channel simply was not a practical option. Kaiser Permanente’s CMT is a prominent example: it combined SNOMED CT with clinician-facing terms, mappings and context-specific subsets as an enterprise terminology resource (Dolin et al. 2004).

Published operational experience shows that this convenience is sustained by ongoing expert work. At the University of Nebraska Medical Center, resolving a problem not yet in its 12,000-term lexicon (finding a code, notifying the clinician and coding the record) took an average of 30 minutes of expert coder time, with unresolved terms escalated to a vocabulary team (Warren et al. 1998). At Hospital Italiano de Buenos Aires, a terminology service built through continuous user feedback recognized 140 ways to describe arterial hypertension, yet 67% of problem-list concepts still required post-coordination, local mappings needed manual work, and hierarchy-based rules had to be reviewed with each release (González Bernaldo de Quirós et al. 2018). At enterprise scale, early CMT tooling had to reconcile conflicting local enhancements from Kaiser Permanente and Mayo Clinic and distribute compatible updates (Campbell et al. 1996). These systems succeed, but their success depends on specialized teams, governance and tooling.

That work creates a substantial maintenance obligation for each implementation. Every additional interface term must be authored, mapped to a concept and kept aligned as the reference terminology evolves; terminology maintenance more generally is complex, resource-intensive and time-consuming, demanding change management, validation, consistency checking and versioning that grow with size (Bakhshi-Raiez et al. 2008). SNOMED International itself notes that a custom interface terminology is “challenging to maintain and keep up to date with new SNOMED versions, with risks of quality issues such as duplication and ambiguity,” and recommends using SNOMED CT’s own descriptions where feasible (SNOMED International 2024). Without sustained upkeep, local interface terminologies and their dependent value sets and mappings drift out of alignment across versions (Bakhshi-Raiez et al. 2008; González Bernaldo de Quirós et al. 2018). This long-recognized tension between flexible, clinician-friendly expression and structured, maintainable representation (Rosenbloom et al. 2011), together with the imbalance between unbounded expressive variety and finite curation capacity, makes an exhaustively curated additional interface vocabulary difficult to sustain.

This frames the question the rest of the paper pursues. If the additional curation could be replaced by retrieval *computed* at query time, the maintenance burden would fall, but only if that computed route can run *beside* the terminology’s own lexical search without degrading it, because otherwise one has merely traded a maintenance problem for an accuracy problem. Reducing local curation and making two retrieval paradigms of opposite nature coexist are therefore the same problem seen from two sides: the motivation and the technical crux.

### Dynamic semantic matching

Recent language technologies offer another way to facilitate search without creating a separate local vocabulary. Biomedical text-representation models place synonyms and paraphrases close in a shared semantic space even without lexical overlap (Remy et al. 2024; Liu et al. 2021); this approach has powered semantic search over large clinical ontologies (notably the CSIRO group’s learned semantic search over SNOMED CT, which outperformed a lexical baseline (Ngo et al. 2021)) and unsupervised concept annotation against SNOMED CT (Abdulnazar et al. 2024). Crucially, semantic matching does not replace the lexical curation performed by SNOMED CT: embeddings are generated from its existing descriptions, so the terminology’s curated lexical content anchors the semantic representation. Combined with LLMs that can normalize and translate clinician shorthand, these methods can dynamically bridge the gap between a natural query and those descriptions, potentially replacing much of the manual expansion with additional local synonyms.

### Two retrieval technologies of opposite nature

These two ways of reaching a concept are of opposite nature in almost every respect. Lexical matching is deterministic, character-level and precise: it does exactly what the user’s keystrokes specify, tolerates partial and out-of-order input, and lets the clinician steer the search interactively, but it cannot cross a wording or language gap for which it was given no shared tokens. Semantic matching is learned, meaning-level and approximate: it crosses paraphrase and language with ease, but it poorly represents abbreviations and half-typed fragments, and cannot be steered keystroke by keystroke. The two are strong and weak on disjoint inputs, and their scores are not even on the same scale. Combining them is therefore not obviously beneficial: for any given query one channel is usually off-task, and a naive combination lets that off-task channel inject distractors that displace the concept the on-task channel has already found. Nor can the difficulty be sidestepped by routing each query to the method that suits it, because in real use the boundary is not stable even within a single user: the same clinician may type a complete term one moment and a two-letter prefix the next, so which regime a query belongs to cannot be read reliably from the query itself, let alone predicted in advance. Delegating that routing decision to a language model on every query (or, in an incremental search box, on every keystroke) would add latency and computational cost out of proportion to the retrieval it is meant to steer. The central design question of this paper is whether two technologies this different can be made to *coexist* in one retrieval flow (each contributing where it is strong, neither degrading the other) without requiring the system to know in advance which regime a query belongs to.

## Objective

We propose a generalizable reference architecture for retrieving SNOMED CT concepts from free-text clinical expressions, and illustrate it with a locally runnable, open-source implementation and a preliminary external evaluation. Its organizing question is how retrieval technologies of opposite nature — deterministic lexical matching and learned semantic matching — can coexist over the same curated descriptions without degrading one another, across five separable functions: query interpretation, complementary lexical and semantic candidate generation, rank fusion, optional re-ranking, and terminology-aware constraints. We do not aim to prescribe a technology stack or a new state-of-the-art entity linker, but to examine *hybrid concept retrieval* as an implementation pattern between direct lexical search and a hand-maintained interface vocabulary — publishing the reference implementation to make its design choices inspectable and reusable.

## Materials and methods

### Reference architecture

At an abstract level, the proposed architecture comprises five separable functions: (1) interpreting or normalizing the input query; (2) generating candidates through complementary lexical and semantic retrieval paths; (3) combining evidence from rankings whose scores may not be directly comparable; (4) optionally re-ranking a small candidate set with a more computationally intensive model; and (5) applying terminology-aware constraints and deterministic safeguards. The architecture does not depend on a particular LLM, embedding model, re-ranker, database or fusion algorithm. Implementers can substitute or omit components according to their languages, latency requirements, available infrastructure and governance constraints. Its central principle is to preserve the precision and responsiveness of lexical matching while adding a semantic route for expressions that do not share the terminology’s wording; and, critically, to combine the two so that neither degrades the other. Functions (3) and (4), fusion and re-ranking, are precisely where that coexistence is secured: fusion admits candidates from whichever channel is on-task, and re-ranking demotes the distractors contributed by the channel that is off-task for a given query. Figure 1 summarizes these functions and the flow between them.

**Figure 1:**
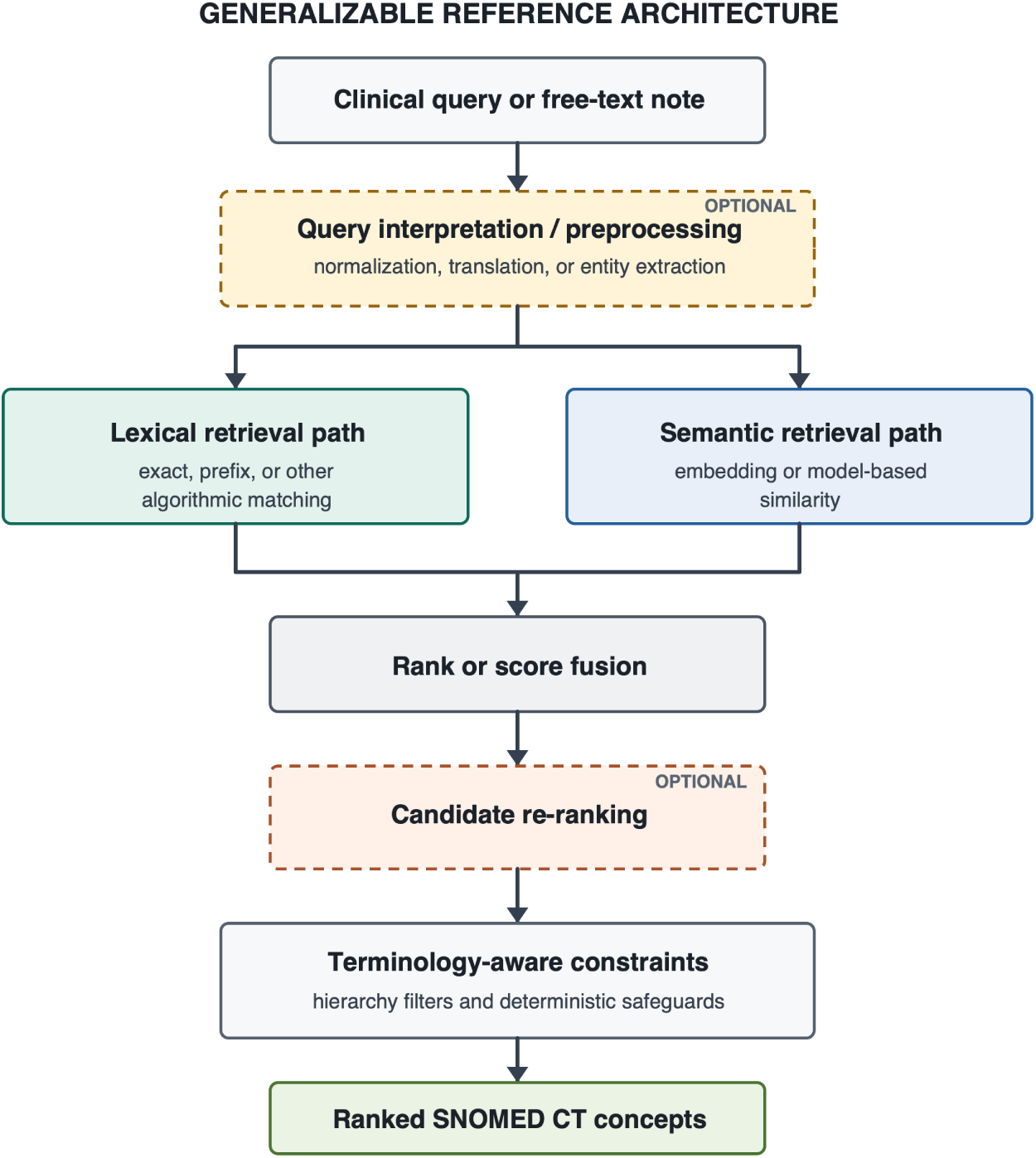
Generalizable reference architecture for hybrid SNOMED CT concept retrieval. Dashed borders indicate optional functions. Components within each function may be substituted according to local requirements; arrows show the flow of queries, candidates and rankings.

The architecture is also language-independent by construction. Its two language-sensitive functions, query interpretation and semantic retrieval, can be instantiated with multilingual models, which already exist off the shelf (multilingual biomedical bi-encoders, LLMs and cross-encoders), so a query in one language is resolved against descriptions in another without any per-language curation: the lexical path stays tied to the language of the indexed descriptions, while the semantic path carries meaning across languages. Cross-lingual retrieval is therefore an available design choice rather than a per-deployment research problem.

### Reference implementation

The system evaluated here is one concrete instance of this model. A local LLM normalizes short queries or extracts clinical entities from longer text; deterministic multi-prefix matching supplies the lexical candidates; a biomedical bi-encoder supplies the semantic candidates; Reciprocal Rank Fusion combines both rankings; an optional cross-encoder re-ranks the fused candidates; and hierarchy filters constrain results to an appropriate SNOMED CT domain. In this implementation, retrieval, fusion and hierarchy filtering are served from a single PostgreSQL instance and executed in one query plan. The following sections describe these choices to make the example reproducible, rather than as requirements of the general architecture. The open-source implementation also provides an executable reference in which components can be inspected, replaced and re-evaluated against different languages, SNOMED CT editions, corpora and infrastructure.

#### Lexical channel

Active descriptions are indexed as PostgreSQL full-text vectors with a custom accent- and case-insensitive, non-stemming configuration and a GIN index. At query time, each fragment entered by the user becomes a prefix clause and the clauses are joined by AND; every fragment must therefore match the initial characters of a word in the description, in any order. For example, *myo inf* is represented as myo:* & inf:* and matches *Myocardial infarction* as readily as *inf myo*. Shared prefixes also absorb some language-dependent inflectional and derivational variation without explicit stemming or lemmatization. This supports incremental search with minimal typing while avoiding the broad result sets produced by matching only some of the entered prefixes. Ranking uses ts_rank with length and unique-word normalization so that concise canonical descriptions outrank verbose ones (PostgreSQL Global Development Group 2026).

#### Semantic channel

Each description is embedded with BioLORD-2023-M, a biomedical, multilingual sentence bi-encoder (Remy et al. 2024) whose lineage traces to self-aligned biomedical representations (Liu et al. 2021), and stored in a pgvector column with an HNSW index for approximate nearest-neighbour search by cosine similarity. For reproducibility we note one index-search setting that materially affects results: the HNSW search width (ef_search) is set equal to the per-channel candidate pool (200) so the index returns the full candidate set, rather than pgvector’s default of 40, which would cap the semantic channel below the requested pool and occasionally return an unrepresentative neighbourhood.

#### Fusion and re-ranking

The two rankings are combined with Reciprocal Rank Fusion (RRF) (Cormack et al. 2009), which requires no score calibration between channels. An optional cross-encoder re-ranker (BGE-reranker-v2-m3, built on a multilingual encoder (Chen et al. 2024)) refines the fused top-*k*; its score is min-max blended with the RRF score rather than allowed to override it. A deterministic rule pins exact-match descriptions to the top.

The re-ranking function admits more than one implementation. Besides the deterministic cross-encoder, which scores surface similarity between the query and each candidate description, we also implement a **contextual LLM selection** (“LLM Pick”): a local LLM is shown the query, the free-text context it appears in, and the numbered top-*k* candidates, and returns the *index* of the best-fitting concept. Because it can only choose from the retrieved list it cannot invent a concept, and its pick is pinned to rank 1. Unlike the cross-encoder it sees the surrounding context the query itself discards, so it resolves homonyms and abbreviations by use (e.g. *MR* as mitral regurgitation in a cardiac context versus an MRI scan when imaging is discussed). It is a second, more expensive instance of the same re-ranking function, applied over the small candidate set.

#### Hierarchy filter

A precomputed transitive closure of the IS-A graph allows any search to be constrained to the descendants-or-self of a concept (e.g. only *Clinical findings*), turning “is concept C under X?” into a single indexed array-containment lookup. This matters because a single surface form often has near-neighbours in several SNOMED CT hierarchies (the same words can name a *finding*, a *morphologic abnormality*, a *body structure* or a *substance*), and constraining the search to the intended domain removes those cross-hierarchy homonyms before they compete for the top rank. In the entity-extraction path the constraint is supplied automatically by the extracted semantic type, so the LLM’s typing decision feeds directly into retrieval precision.

#### LLM roles

A local LLM (gemma 4 (Gemma Team, Google DeepMind 2026), a reasoning model run with its chain-of-thought disabled for direct output, served through an OpenAI-compatible endpoint) plays two roles. (1) *Query normalization*: it translates a short clinician query from any language to one canonical English clinical term and expands abbreviations, so that both retrieval channels operate on clean English; the embedding model’s crosslingual nature also allows the system to degrade gracefully without the LLM. (2) *Entity extraction*: given a free-text note, it returns structured entities as JSON, including the verbatim span, semantic type, assertion/temporal context (present/absent/past/unknown), laterality, severity and an English clinicalTerm. Each extracted entity is then mapped independently through the hybrid engine, using its normalized clinicalTerm as the query and its semantic type as the hierarchy filter (falling back to an unfiltered search when the type filter would exclude everything).

### Data and implementation

We used the RF2 Snapshot view of the 1 June 2026 SNOMED CT International Edition production release. We loaded the active English-language descriptions of active concepts: 1,024,825 descriptions (381,856 fully specified names and 642,969 synonyms) for 381,856 concepts; of these, 1,020,940 unique normalized description strings were embedded. A transitive closure of the ISA relationships was precomputed over all 381,856 active concepts to support the hierarchy (descendant) filter. The stack runs locally on a single workstation (Apple Silicon), with the database in a container and all model inference in-process; no clinical data leaves the machine. The full pipeline, indexes and demonstration interface are reproducible from a public repository (https://github.com/alopezo/snomed-hybrid-search).

### External evaluation

#### Choice of corpus

Openly licensed gold standards that link *free clinical text* to SNOMED CT are scarce. Most English clinical-note resources that normalize to SNOMED CT or the UMLS require a data-use agreement, and general clinical NLP corpora do not target SNOMED CT. The Spanish corpora produced by the Barcelona Supercomputing Center for the BioASQ/BioCreative shared tasks are a notable exception: they are released under CC BY 4.0 and every mention is normalized to SNOMED CT. We evaluated on **DisTEMIST** (BioASQ 2022), 1,000 Spanish clinical case reports with disease mentions linked to SNOMED CT (Miranda-Escalada et al. 2022) — a 750/250 train/test split, of which we use the 750-document training set — because diseases map cleanly onto the *Clinical finding* hierarchy. Two caveats follow from this choice and we treat them explicitly rather than hide them. First, the corpus is **Spanish**; for a multilingual system this is less a limitation than a cross-lingual stress test, since the query and the terminology descriptions are in different languages. Second, the texts are drawn from **published case reports**, whose language is more edited than bedside notes; the evaluation therefore probes concept normalization more than the full messiness of real electronic health record text. We use this corpus because it is among the few open, SNOMED-CT-linked options, and read the results with these limits in mind.

To probe exactly those two limits, language and real-EHR messiness, we added a second corpus: the **SNOMED CT Entity Linking Challenge** (DrivenData / PhysioNet, 2024; henceforth the **EL Challenge**), whose released training set is 272 English MIMIC-IV-Note discharge summaries exhaustively annotated with 74,808 mentions linked to SNOMED CT across 6,624 distinct concepts (Davidson et al. 2025) (its evaluation test set is held out on DrivenData, so, as with DisTEMIST, we evaluate on the training split). It is English and drawn from real electronic health records, and it spans the full concept range (findings, procedures, body structures, and more) rather than diseases alone. Access is credentialed through PhysioNet under the MIMIC data-use agreement, which precludes redistribution, so we use it locally and do not share it. The two corpora bracket the target regime: DisTEMIST (Spanish, edited case reports, disease-scoped) and the EL Challenge (English, real EHR, all clinical domains).

#### Task and configurations

To measure the *search* capability independently of our entity extractor, we ran a **search-only** evaluation: each gold mention’s verbatim span is submitted as a query and we record the rank of the gold SNOMED CT code; this is the corpus’s own entity-linking (normalization) subtask. We swept the two learned, optional components of the search pipeline as a 2×2 design, LLM query pre-processing (gemma normalization/translation) on/off × cross-encoder re-ranking on/off, and report three of the four cells. The lexical and semantic channels and their RRF fusion were always active; the hierarchy filter was fixed to *Clinical finding* (the corpus is diseases); and we retrieved *k* = 10 candidates.

#### Edition-drift resolution

The gold was annotated against a circa-2022 edition; by our loaded release (International 2026-06-01) some gold concepts had been inactivated. Because a fixed gold standard drifts as the terminology evolves (itself an instance of the maintenance problem this paper is about), we resolved every gold code through SNOMED CT’s **historical-association** reference set (in priority order SAME AS → REPLACED BY → POSSIBLY REPLACED BY → POSSIBLY

EQUIVALENT TO → ALTERNATIVE) to its current active concept before scoring.

#### Scope

The system targets *disorder* and *finding* concepts. Where the annotators coded a mention to a *substance* or *morphologic-abnormality* concept for the same clinical entity (for example, *renal stone* as a substance rather than as a disorder), a system that returns the disorder can never match by identifier. We therefore scored only gold whose resolved concept is a disorder or finding, so that representation choices outside the system’s target do not create artificial misses.

#### Metrics

We chose metrics that mirror how a clinician uses a concept picker. In a typical search interface the user types a term and selects from a short ranked list, so what matters is whether the intended concept appears *near the top*. We therefore report, under a **strict** rule (our concept identifier equals the resolved gold): **accuracy@1**: the top candidate is exactly the gold concept; **recall@5** and **recall@10**: the gold concept is among the first 5 or 10 candidates, i.e. the options a user would see in a drop-down list without, or with a small scroll; and **mean reciprocal rank (MRR)**, which rewards a higher position of the gold concept. We deliberately foreground recall@5 and recall@10 because they answer the practical question for a picker interface, “would the right concept be on the screen for the user to choose?”, rather than requiring the system to be right on its single best guess. Separately, because a *parent* or *child* of the intended concept can be an acceptable or navigable alternative, we report a **near-miss** rate: the additional fraction of mentions (beyond the exact matches) whose topk list contains an ancestor or descendant of the gold. The near-miss rate is thus an increment on strict recall, not a replacement for it: the two together sum to a “same-lineage” recall. It is an upper bound on “clinically acceptable” rather than a hard accuracy, since a hierarchical neighbour is not always an acceptable substitute; confirming that would require clinician adjudication.

#### Field-scoped queries and context-aware pre-processing

The EL Challenge corpus lets us model how these concepts are entered in practice. Real clinical data-entry interfaces (typeahead boxes, drop-downs) are *scoped by field*: a diagnosis field searches findings, a procedure field searches procedures, a medication field searches products. We simulate this by constraining each query to the top-level SNOMED CT hierarchy of its own gold concept (the field’s domain), rather than to a single fixed hierarchy. The domain is a large hierarchy (of order 10^5 concepts) that stands in for the entry field the clinician chose; it is application context, not the answer, and every query in a field receives the same constraint blind to its target. This estimates what an implementer of a field-scoped picker would obtain. Real EHR text is also dense in ambiguous shorthand (for example *MMM*, *CTAB*, *RRR*) that the pre-processing LLM cannot disambiguate from the isolated span. We therefore also tested a **context-aware pre-process**: a single LLM call that normalizes the mention given the surrounding note chunk rather than the span alone.

## Results

### Feasibility of the reference architecture

The open-source reference implementation instantiated all five functions of the proposed architecture in an end-to-end pipeline. For short expressions, it generated lexical and semantic candidates and returned a fused ranking. For longer notes, it first extracted and normalized clinical entities and then retrieved candidates independently for each entity. Optional re-ranking and hierarchy constraints operated on the same candidate flow, allowing the architecture to support both interactive concept search and note-level entity linking. The same open-source application, an interactive search interface, a note-level entity-extraction view and a batch evaluation harness, was the instrument for both the qualitative observations and the quantitative results that follow. This establishes the technical feasibility of the architectural pattern; the observations and metrics below characterize this particular implementation.

### Illustrative component behaviour

The examples exposed complementary roles for the components rather than a consistent advantage for any single model. The semantic path recovered lay phrasing with no shared tokens (“water on the knee” → *Effusion of knee joint*), whereas deterministic multi-prefix matching retrieved a clinician abbreviation (“inf myo” → *Myocardial infarction*) that was represented poorly by the embedding model. Query interpretation and cross-lingual retrieval supported inputs such as “radiografía de tórax” → *Plain X-ray of chest* and “EPOC” → *Chronic obstructive pulmonary disease*. For longer notes, the LLM extracted entities and represented assertion separately from concept identity, mapping “no fever” to *Fever* with an absent flag and “history of stroke” to *Cerebrovascular accident* with a past flag. In a Spanish note, entities were extracted verbatim, normalized to English clinicalTerms, and mapped to the expected SNOMED CT concepts (e.g. *diabetes mellitus tipo 2* → *Diabetes mellitus type 2*; *disnea de esfuerzo* → *Dyspnea on exertion*).

### External evaluation on DisTEMIST

We evaluated all DisTEMIST training documents, which contain 1,057 unique disease mentions; 37 gold codes were recovered through historical associations, while 66 mentions were scoped out as non-disorder/finding and 6 could not be resolved to an active concept in our release, leaving **985** mentions. Table 1 reports the three search configurations.

**Table 1:** Search-only linking on 985 DisTEMIST disease mentions (Spanish, zero-shot). *PP* = LLM query pre-processing (normalization); *RR* = cross-encoder reranking. *MRR* = mean reciprocal rank of the gold concept. The strict columns *acc@1*, *R@5* and *R@10* (accuracy@1, recall@5, recall@10) require the exact concept identifier and read as a picker list: is the gold concept on the screen the user would see? *+near@10* is the additional fraction whose top-10 held an ancestor or descendant of the gold (the near-miss increment), and *=lin@10* is the two summed (same-lineage@10): the most optimistic reading, in which any hierarchical neighbour counts as acceptable. Best value per column in **bold**.

| PP | RR | MRR | acc@1 | R@5 | R@10 | +near@10 | =lin@10 |
| --- | --- | --- | --- | --- | --- | --- | --- |
| off | **on** | **0.65** | **0.57** | **0.77** | **0.80** | +0.10 | **0.90** |
| on | off | 0.63 | 0.56 | 0.71 | 0.77 | +0.11 | 0.88 |
| on | on | 0.64 | 0.57 | 0.72 | 0.77 | +0.11 | 0.88 |

Read the numbers as a concept picker: recall@5 and recall@10 are the chance that the intended concept is on the short list a clinician would scan and select from, while accuracy@1 is the chance the system’s first candidate is already correct. Three findings stand out. First, the **cheapest configuration is also the best**: with query pre-processing *off* and re-ranking *on*, the exact concept is the top candidate for 57% of mentions (accuracy@1 = 0.57) and appears within the top-10 picker list for 80% (recall@10 = 0.80); for a further 10% of mentions a parent or child of the intended concept is in that list (near-miss@10 = +0.10). Because this configuration makes no per-query LLM call, it is simultaneously the most accurate and the least expensive to run. Second, **LLM query pre-processing still slightly hurts on this task**: enabling it lowers accuracy@1 from 0.57 to 0.56 (a smaller penalty than with the previous-generation LLM). The gold mentions are already clean, terminology-like disease terms, and the multilingual embedding maps them cross-lingually without help; the LLM’s normalization to English instead shifts the semantic neighbourhood (for example, *infertilidad* → *infertility* pulls the specific “X infertility” disorders above the exact generic *Infertile*). The LLM’s value is for the dirty, lay or abbreviated input of interactive use, not for well-formed terminology strings. Third, **reranking helps** when it is not preceded by aggressive normalization, improving both accuracy@1 and MRR; the two components are genuinely optional and the optimal configuration is task-dependent.

The **MRR of 0.65** is only 0.08 above accuracy@1 (0.57), and that small gap is the informative part: of the mentions the system retrieves, almost all sit at or just below the top rather than buried deep in the list. Decomposing the 985 mentions, roughly 57% are exact at rank 1, about 23% are retrieved at a lower rank (around rank 3), and the remaining 20% are not retrieved within the disorder/finding scope at all. The residual difficulty is therefore one of *recall* (the concepts the system never surfaces) rather than of *ranking*: when the intended concept is present, the picker rarely makes the clinician scroll for it. The columns bound the outcome from both sides: strict accuracy@1 = 0.57 is the pessimistic floor, while same-lineage@10 = 0.90 is the optimistic ceiling in which any hierarchical neighbour counts as acceptable, with the true operational value depending on how a given workflow treats near-misses.

These are **zero-shot** results (no training or fine-tuning on the corpus). They are not directly comparable to the results reported for the DisTEMIST shared task: its best supervised linking system reached a micro-averaged F1 of 0.57 (Miranda-Escalada et al. 2022), but that figure is *end-to-end*, scoring mention detection and normalization together, whereas our search-only accuracy is computed on gold mention spans and isolates the normalization step that is this paper’s subject. The two are of similar magnitude but measure different tasks; a like-for-like comparison would require scoring our full extraction-and-mapping pipeline end-to-end, which (as the present study concerns the search step rather than entity extraction) we leave to future work. The near-miss rate shows that a meaningful share of the strict “errors” are hierarchically adjacent rather than wrong: at rank 1, beyond the 0.57 exact matches, a further 0.16 of mentions return a parent or child concept as the top candidate. Whether such a neighbour is acceptable is a clinical judgement, so we treat this as an upper bound rather than as accuracy. A further residual class, credited by neither the exact nor the near-miss count, is the annotators’ choice of a *substance* or *morphologic-abnormality* concept where the system returns the corresponding *disorder*, a genuine representation difference in the gold itself, not a retrieval failure.

Running the full extraction-then-mapping pipeline over the same notes (rather than the gold spans) lowers these figures, which isolates the contribution of the extraction stage: the LLM occasionally mis-types or omits an entity, and long notes must be chunked to prevent the small model from degenerating and dropping their tail. In this implementation the search is the stronger component; extraction is where accuracy is currently lost.

### Coexistence of the two retrieval channels

To test whether the two channels coexist without a trade-off, we repeated the best serving configuration (no query pre-processing, re-ranking on) while restricting retrieval to a single channel (Table 2). The semantic channel accounts for essentially all of the accuracy on this corpus, and fusing the lexical channel back in leaves it unchanged (accuracy@1 0.57 vs 0.58; the difference is a single mention). The lexical channel alone collapses (accuracy@1 0.06) for a specific reason: the queries are Spanish while the indexed descriptions are English, so without the query-normalization step the lexical channel has almost no tokens to match, whereas the multilingual embedding bridges the two languages directly. The informative result is the near-identity of the semantic and fused rows: on clean cross-lingual normalization, adding the lexical channel neither helps nor harms.

**Table 2:** Retrieval-channel ablation on the same 985 DisTEMIST mentions, in the best serving configuration (no query pre-processing, re-ranking on); only the retrieval channel varies. *both* is the Reciprocal Rank Fusion of the two channels.

| channel | acc@1 | recall@10 | MRR |
| --- | --- | --- | --- |
| lexical only | 0.06 | 0.06 | 0.06 |
| semantic only | **0.58** | **0.80** | **0.66** |
| lexical + semantic (fused) | 0.57 | 0.80 | 0.65 |

### Contribution of the hierarchy filter

The hierarchy filter, fixed to *Clinical finding* throughout, contributes at the top of the ranking. Repeating the best configuration without it lowers accuracy@1 from 0.57 to 0.53 and MRR from 0.65 to 0.62, while recall@10 barely moves (0.80 to 0.78). The pattern is that of a precision tool: the correct concept is usually retrieved within the top-10 either way, but without the domain constraint, concepts from other SNOMED CT hierarchies (a *substance*, a *morphologic abnormality* or a *body structure* sharing the query’s words) sit above it and displace it from rank 1. Because the constraint is a subtree test over a precomputed transitive closure, this precision gain costs a single indexed lookup.

### English, real-EHR validation on the SNOMED CT Entity Linking Challenge

On the EL Challenge corpus (English MIMIC-IV discharge notes, all concept types) we ran the interactive serving configuration (no query pre-processing, re-ranking on) with each query scoped to its own domain, simulating a field-scoped typeahead over the 12,897 unique (span, concept) mentions that remain after collapsing spans repeated across notes and keeping the single-concept annotations (from the corpus’s 74,808 total). Overall accuracy@1 was 0.52 and recall@10 0.73 (Table 3): a clinician would find the intended concept on the top-10 picker list about three times in four. Stratified by domain, the *finding* stratum, directly comparable to DisTEMIST, reached accuracy@1 0.54 and recall@10 0.75, a little below DisTEMIST’s 0.57 and 0.80, consistent with real EHR text being messier than edited case reports; procedures were hardest (accuracy@1 0.45) and body structures close to findings (0.54). Once each field is scoped, performance is thus broadly even across clinical domains rather than confined to diagnoses.

**Table 3:** Field-scoped linking on the SNOMED CT Entity Linking Challenge (English MIMIC-IV notes; no query pre-processing, re-ranking on), stratified by the gold concept’s domain. Each query is constrained to its own domain’s top-level hierarchy, simulating a scoped data-entry field. *R@5*/*R@10* = recall@5/recall@10; *+near@10* = near-miss increment (top-10 holds a parent/child).

| domain | n | acc@1 | R@5 | R@10 | MRR | +near@10 |
| --- | --- | --- | --- | --- | --- | --- |
| finding | 7,318 | 0.54 | 0.71 | 0.75 | 0.61 | +0.08 |
| procedure | 3,073 | 0.45 | 0.63 | 0.69 | 0.53 | +0.12 |
| body<br>struct. | 2,491 | 0.54 | 0.70 | 0.74 | 0.60 | +0.10 |
| **all** | **12,897** | **0.52** | **0.69** | **0.73** | **0.59** | +0.09 |

A discrepancy analysis tempers the strict accuracy. Of the 6,186 mentions not exact at rank 1, 21% place the gold concept within the top-5 or top-10 (still on the picker list) and a further 9.4% return a hierarchical neighbour; only 17% are hard misses. Inspecting the hard misses, roughly a quarter are dense clinical abbreviations and physical-exam shorthand (*AOx3*, *HEENT*, *CTAB*, *RRR*, *EOMI*) that neither retrieval channel can resolve from the isolated span: the lexical channel matches the wrong fragment (*NAD* → *Nadolol*), and the LLM, normalizing the bare abbreviation, hallucinates (*MMM* → *myocardial infarction*, a clinically unsafe expansion of *moist mucous membranes*). Part of this class is also a *scope* difference rather than an error: the EL Challenge annotates normal exam findings exhaustively, whereas a clinical extractor typically omits them.

These abbreviations are precisely where local context is decisive. On a sample of 250 hard misses, a **context-aware pre-process** (a single LLM call that normalizes the mention given its note chunk) recovered the gold concept within the top-10 for **48% of them (accuracy@1 28%)**, against **26% (accuracy@1 15%)** for the same LLM applied to the bare span: context roughly doubled recovery and, at rank 1, nearly doubled it. It disambiguated shorthand (*EOMI* → *extraocular movements intact*, *CTAB* → *clear to auscultation bilaterally*, *HI* → *homicidal ideation*), fixed typos (*bnzodzpn* → *benzodiazepine*), and resolved context-dependent tokens (*LOW* → *low platelet count*), while removing the unsafe isolated-span hallucinations; and it needs no change to the entity extractor. Applied to *every* mention across the whole corpus, however, context-aware preprocessing did not raise the aggregate: strict accuracy@1 fell from 0.52 to 0.49 while the same-lineage rate rose slightly (hier@10 0.83 to 0.84, near-miss +0.09 to +0.12). The reason mirrors the DisTEMIST finding: most mentions are already resolved by the bare-span search, and re-normalizing those clean terms perturbs some of them out of rank 1, a loss on the easy majority that slightly outweighs the rescue of the hard minority. Context-aware pre-processing is therefore best used *selectively*, as a second-pass rescue when the direct search fails or returns a low-confidence result, rather than as a mandatory first step (the same fallback logic already used for the hierarchy filter). Together the two corpora give a consistent account of when LLM normalization helps: it *hurts* clean, terminology-like input (DisTEMIST, and the easy majority of the EL Challenge) but *rescues* the messy shorthand of real EHR (the EL Challenge’s hard misses), most of all when it can see the surrounding context.

### Contextual re-ranking with an LLM

Re-ranking need not be a fixed model. We evaluated the contextual LLM selection described in *Fusion and re-ranking* as an alternative implementation of the re-ranking function, layered over the best serving configuration on the full corpora (Table 4). It is distinct from the context-aware pre-processing above: that rewrites the *query* before retrieval and is free to drift or hallucinate, whereas this reorders the *already-retrieved* candidates and can only pick from them, so it cannot invent a concept. On both corpora it lifted top-rank accuracy — DisTEMIST accuracy@1 0.57→0.61 (MRR 0.65→0.68) and the EL Challenge 0.52→0.56 (MRR 0.59→0.62) — while, because it only reorders the retrieved list, recall@5 and recall@10 were unchanged: the gain is concentrated at rank 1, exactly where a picker is judged. The hierarchy-aware figures rose in step, so the reordering moves same-lineage concepts up rather than trading correctness for neighbours. This is the mirror image of context-aware pre-processing, which, being free to rewrite the query, lowered the aggregate on the same corpus: grounding the LLM to the retrieved candidate set is what turns its contextual judgement from a liability into a gain.

**Table 4:**
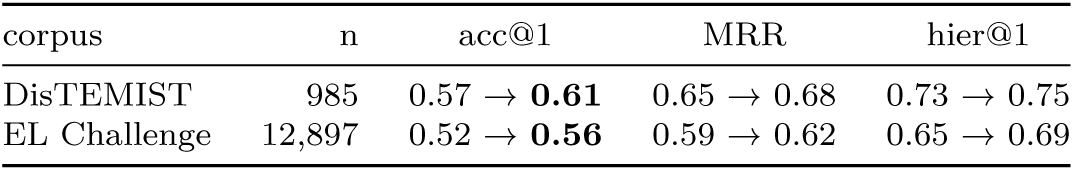
Contextual re-ranking (“LLM Pick”) as an alternative implementation of the re-ranking function, over the best serving configuration (no query pre- processing, cross-encoder re-ranking on) on the full corpora. The layer reorders the top-*k* only, so recall is unchanged; each cell shows *baseline* → *with LLM Pick*.

| corpus | n | acc@1 | MRR | hier@1 |
| --- | --- | --- | --- | --- |
| DisTEMIST | 985 | 0.57 → **0.61** | 0.65 → 0.68 | 0.73 → 0.75 |
| EL Challenge | 12,897 | 0.52 → **0.56** | 0.59 → 0.62 | 0.65 → 0.69 |

Two implementation details mattered. First, the context must be light: a generic framing (“clinical data entry in an EHR”) worked best, while feeding the mention’s full surrounding sentence *lowered* accuracy on the EL Challenge, because the messy note text led the model to over-interpret a clean lab or finding into a more specific concept than the gold. Second, a one-line hint encoding a SNOMED CT naming convention — anatomy is *Structure of X* by default, *Entire X* only for the whole organ — was decisive on the EL Challenge: without it the model systematically picked the rarer *Entire X* concept for body sites and the layer was net-negative; with it, it became net-positive. The layer is roughly fifteen to twenty times slower than the deterministic cross-encoder (one local-LLM call per query), so, like query pre-processing, it is best applied selectively rather than to every query.

## Discussion

### Principal findings

This study contributes a generalizable reference architecture and an executable, open-source implementation. The implementation demonstrates the technical feasibility of combining query interpretation, complementary lexical and semantic retrieval, rank fusion, optional re-ranking and terminology-aware constraints in a single concept retrieval flow. Illustrative examples showed that these functions address different forms of vocabulary mismatch, while the preliminary external evaluation established a reproducible baseline for one concrete configuration. A more specific finding sharpens the architectural claim: two retrieval technologies of opposite nature can be fused without a trade-off, provided re-ranking suppresses whichever channel is off-task for a given query (developed below). A recurring design principle runs through the evaluation: the expensive, optional components pay off when applied *conditionally* rather than uniformly (re-ranking only over the small fused candidate set, the hierarchy filter with a fallback to unconstrained search, and LLM preprocessing as a selective rescue for input the direct search cannot resolve), because applied to every query they can degrade the easy majority that the cheap deterministic and semantic paths already handle. These findings support the coherence and feasibility of the architectural pattern. They do not establish that the selected components are optimal, that the observed performance transfers to other implementations, or that the system is ready for clinical use.

#### Implications for terminology infrastructure

Hybrid concept retrieval provides an intermediate strategy between expecting clinicians to resolve every mismatch through lexical query refinement and attempting to anticipate every likely expression in an additional interface vocabulary. A responsive lexical path preserves familiar, precise interaction with SNOMED CT descriptions, while a semantic path computes further candidates when the query uses different wording. Both paths remain anchored in lexical content curated and distributed with SNOMED CT. This may reduce local term-by-term authoring and mapping, but it does not make maintenance disappear. Instead, it shifts work toward governing models, indexes, constraints, terminology releases and regression tests. Some of this work can be automated and shared, whereas exhaustive local phrase curation grows with the variety of clinician language (Furnas et al. 1987). When the retrieval models are multilingual, as in our implementation, that computed bridge spans languages as well, so a deployment need not curate colloquial expressions language by language. Additional curated interfaces remain appropriate when highly guided workflows, local policy or limited opportunities for user training make them valuable. Using SNOMED CT’s own descriptions directly where feasible also follows SNOMED International guidance (SNOMED International 2024).

#### Coexistence without a trade-off

The channel ablation (Table 2) shows the semantic channel carries clean disease normalization on DisTEMIST, and that fusing the lexical channel back in costs nothing measurable. Read naively this argues for dropping the lexical path, but that reading is an artifact of the benchmark: DisTEMIST mentions are complete, edited disease terms that never exercise what the lexical channel is uniquely good at, order-independent matching of partial, as-typed input such as “inf myo” for *Myocardial infarction*, a fragment the embedding represents poorly. The two channels serve different regimes, and a single query does not announce which one it belongs to. Rank fusion followed by cross-encoder re-ranking is what lets one pipeline keep both: fusion contributes candidates from whichever channel is on-task, and the reranker suppresses the off-task one so it does not degrade the result. Naive equal-weight fusion *without* re-ranking does lose ground to the stronger single channel, whereas re-ranking restores parity. The payoff is regime-robustness without query routing: the same system serves a Spanish disease term and a half-typed English abbreviation, without classifying the query in advance. The benchmark exercises only one regime, so it should be read as evidence that hybridization is *safe* for clean normalization, not that the lexical channel is dispensable.

These two regimes map onto the two modes of clinical data entry noted at the outset, and the mapping suggests that the value of hybridization is not uniform across callers. An automated agent typically submits a complete, already-normalized term (precisely the input the semantic channel handles best), so for agent-driven encoding the semantic path alone may be sufficient, and the lexical channel adds little. A clinician entering data interactively types partial, abbreviated, as-typed fragments and refines them keystroke by keystroke: the regime in which the lexical channel is indispensable and in which the hybrid most improves usability. Hybridization therefore earns its keep chiefly at the human, interactive end of the spectrum, while a well-formed agent query may be served by semantics alone, though the same pipeline serves both without having to know which is calling.

#### The hierarchy filter as a cheap precision lever

Constraining a search to the expected hierarchy raised top-rank precision at negligible cost (accuracy@1 +0.05, MRR +0.03) by removing homonyms drawn from unintended hierarchies (Section *Contribution of the hierarchy filter*). This puts the terminology’s *structure*, not only its curated descriptions, to work, and makes the LLM extractor’s (or an automated agent’s) semantic-type classification a direct lever on precision: a correct type narrows retrieval to the right domain for the price of one indexed subtree test. A wrong type can exclude the intended concept, a recall risk the implementation mitigates by falling back to an unconstrained search when the filtered query returns nothing.

#### Architecture, implementation and validation

The five architectural functions define separable responsibilities rather than fixed products. Implementations may use different language models, embedding models, lexical algorithms, fusion methods, databases and constraints, and their results may therefore differ substantially. The re-ranking function illustrates this concretely: we implemented it two ways — a deterministic cross-encoder and a contextual LLM selection — and the second improved top-rank accuracy on both corpora (Table 4), an alternative instance of the same function rather than a new one. Performance claims should attach to a specified, versioned implementation, while support for the reference architecture should accumulate across implementations and settings. The open-source system and evaluation materials provide a starting point for a reusable evaluation harness: canonical queries, external corpora, ranking metrics, latency measures and failure cases can be rerun when a component changes. Automated regression testing can make iterative improvement less costly, but it cannot replace validation against the intended users and clinical use case. The required depth of that validation should reflect the consequences of an incorrect match and whether the output is advisory, user-confirmed or recorded without review.

### Relation to prior work

Semantic search over clinical ontologies and SapBERT/BioLORD-style bi-encoders established that embeddings can recover concepts under vocabulary mismatch (Ngo et al. 2021; Liu et al. 2021; Remy et al. 2024; Abdulnazar et al. 2024), while recent scoping work catalogues uses of LLMs with SNOMED CT (Chang and Sung 2024). The distinctive contribution here is architectural rather than a new retrieval model. The closest precedent, the CSIRO system, benchmarked a learned semantic model against a lexical baseline and found the semantic model superior (Ngo et al. 2021); our channel ablation replicates that ordering on a clean-normalization corpus. Where that line of work asks *which single method retrieves best*, we ask *how* a deterministic lexical method and a learned semantic one can coexist in one flow, so that the strength of each is available without the channel that is wrong for a given query degrading the result. The design assigns bounded roles to an LLM for query interpretation and entity extraction, preserves a deterministic lexical path, adds semantic candidate generation, combines rankings without requiring score equivalence, permits more expensive re-ranking only over a small candidate set, and applies constraints derived from the terminology. It connects these technical choices to the operational question of whether access to SNOMED CT can be made more forgiving without creating another large vocabulary to maintain.

### Limitations

The external evaluation covers two corpora (Spanish disease case reports and English real-EHR discharge notes) and the search-only linking task on gold mention spans; a full end-to-end extraction evaluation, other languages, and other note genres remain to be tested, and the context-aware pre-process was quantified only on a sample of hard misses, not integrated and run over a whole corpus. Both gold standards annotate mentions our target extractor deliberately omits (normal exam findings, lab values), so the strict figures are a lower bound on the concepts the system is built to code. The strict/hierarchy-aware gap indicates that the exact-identifier metric understates semantically acceptable retrieval, but neither metric was confirmed by blinded clinical adjudication. We ablated the two optional components (query pre-processing and re-ranking), the hierarchy filter, and the two retrieval channels in isolation, but did not substitute the embedding model, lexical algorithm or fusion method, so the contribution of those functions is not estimated. We also did not compare clinician search performance, measure longitudinal maintenance effort, or evaluate safety in a production workflow. LLM entity typing remains imperfect, extraction latency is LLM-bound, and mapping to a single best concept omits post-coordination. Open source makes the design choices transparent and facilitates independent evaluation, but does not remove the need to validate each implementation against its intended users, terminology edition, language and use case. Next steps include larger evaluations across findings, diseases and procedures; component substitution and ablation experiments; hierarchy-aware and clinician-adjudicated metrics; usability studies; an evaluation on interactive, partially-typed input that exercises the lexical channel’s regime, which a corpus of complete disease terms does not; learned or weighted rank fusion, since equal-weight fusion proved sub-optimal when one channel is off-task; and longitudinal measurement of maintenance work.

## Conclusion

SNOMED CT’s curated lexical content can serve as the foundation for an interface that is partly *computed* at query time. The proposed reference architecture offers a testable, implementation-independent path for combining lexical precision with semantic flexibility so that two technologies of opposite nature coexist over the same descriptions without degrading each other, a coexistence secured by rank fusion and re-ranking rather than by choosing between the paradigms. Its open-source implementation demonstrates feasibility and enables adaptation and re-evaluation, but each resulting system requires validation in its intended setting. The benefit of that coexistence appears to fall unevenly on the two forms of clinical data entry: an automated agent, which submits well-formed normalized terms, may be served by the semantic path alone, whereas an interactive human, who types partial and abbreviated fragments, gains the most usability from the hybrid. This approach may reduce additional local lexical curation, while hand-built interface terminologies retain a role in guided, low-friction clinical workflows.

## Data Availability

All data produced are available online at https://github.com/alopezo/snomed-hybrid-search

https://physionet.org/content/snomed-ct-entity-linking-challenge/

https://zenodo.org/records/6532684

## Acknowledgements

AI coding and writing assistants were used during this work. OpenAI Codex and Claude Code (Anthropic) were both used during manuscript preparation to assist with language editing, structural revision and drafting; Claude Code was additionally used as a programming assistant to help develop the reference implementation, tooling and evaluation frameworks. All AI-assisted content and code was critically reviewed, verified and revised by the authors, who take full responsibility for the final manuscript.

## Ethics

This study did not involve any interaction with human subjects or collection of new data by the authors. It analysed only two previously collected, de-identified, human-derived corpora under their respective licences and data-use agreements. DisTEMIST (Miranda-Escalada et al. 2022) is a corpus of Spanish clinical case reports, openly available under a CC BY 4.0 licence prior to the initiation of this study. The SNOMED CT Entity Linking Challenge dataset (Davidson et al. 2025) comprises de-identified English MIMIC-IV-Note discharge summaries obtained under PhysioNet credentialed access and the MIMIC data-use agreement, which required registration, completion of the required training, and signing a data-use agreement; these data were used locally under that agreement and are not redistributed. MIMIC-IV is fully de-identified in accordance with HIPAA Safe Harbor, and its collection was approved by the Institutional Review Board of the Beth Israel Deaconess Medical Center with a waiver of informed consent. No additional ethics approval was required, as this work used only previously collected, de-identified datasets under their existing licences and agreements.

## Data and code availability

The reference implementation, configuration and evaluation materials are available as open-source software under the Apache License 2.0 at https://github.com/alopezo/snomed-hybrid-search. They are provided to make the architectural choices inspectable and to support adaptation and re-evaluation in other environments. DisTEMIST (Miranda-Escalada et al. 2022) is available under CC BY 4.0 from its authors (https://zenodo.org/records/6532684) and is not redistributed here. The SNOMED CT Entity Linking Challenge dataset (Davidson et al. 2025) is available from PhysioNet under credentialed access and the MIMIC data-use agreement (https://physionet.org/content/snomed-ct-entity-linking-challenge/), which precludes redistribution; it is used locally and not shared here.

